# Rapid growth in autism spectrum disorder referrals reshaped rehabilitation service use: A longitudinal cohort study of children and adolescents in Brazil

**DOI:** 10.64898/2026.08.04.26359668

**Authors:** Juan P. Aguilar Ticona, Ana F. Ferreira-Stagliorio, Gustavo N. de Oliveira Costa, Luciana Moreira, Arthur S. B. Dias, Camila Costa, Ariana Oliveira Santos, André A. de Queiroz, Raul R. de Oliveira Pacheco, Iris Montaño-Castellón, María B. Arriaga, Eduardo M. Netto

## Abstract

**Background:** The global increase in autism spectrum disorder (ASD) diagnoses is expected to substantially increase demand for long-term rehabilitation services. However, little is known about how this increase affects rehabilitation service utilization and capacity in low- and middle-income countries.

**Methods:** We conducted a retrospective longitudinal study of children receiving developmental care at a tertiary rehabilitation center in Salvador, Brazil (2017– 2026). Patients were classified into Childhood Autism, Other ASD, and non-ASD diagnostic groups according to ICD-10 diagnoses. Temporal trends in admissions and patients under follow-up were analyzed using generalized additive models and segmented Poisson regression. Factors associated with follow-up duration were evaluated using multivariable Cox proportional hazards models.

**Results:** Among 2,123 eligible children, 833 (39.2%) had Childhood Autism, 462 (21.8%) had Other ASD, and 828 (39.0%) had non-ASD diagnoses. Compared with children with non-ASD diagnoses, those with Childhood Autism entered care at younger ages, were predominantly male (77.9% vs. 57.2%), attended more visits, and remained under follow-up longer (all *P*<0.001). Admissions of children with Childhood Autism increased by 30.2% annually before 2023 but declined thereafter (−19.6% annually; *P*<0.001). Despite this decline, the number of children with Childhood Autism receiving ongoing rehabilitation continued to increase, reflecting prolonged follow-up. In adjusted analyses, Childhood Autism was associated with a substantially lower hazard of reaching the last recorded follow-up visit than non-ASD diagnoses (adjusted hazard ratio, 0.35; 95% CI, 0.30–0.40; *P*<0.001).

**Conclusions:** The rapid increase in ASD admissions fundamentally reshaped rehabilitation service utilization. Because children with ASD remained under follow-up substantially longer than those with other developmental conditions, they accounted for an increasing share of the rehabilitation caseload, even after new admissions began to decline. These findings highlight the importance of planning rehabilitation services according to both new admissions and the cumulative demand generated by long-term follow-up.

## Introduction

Autism spectrum disorder (ASD) is a neurodevelopmental condition characterized by persistent difficulties in social communication and interaction, together with restricted and repetitive patterns of behavior, interests, or activities that emerge early in life [1]. Although historically considered an uncommon condition, ASD is now one of the most frequently diagnosed neurodevelopmental disorders in childhood [2]. Global prevalence is estimated at approximately 1%, with substantial geographic variation reflecting differences in awareness, diagnostic practices, and surveillance systems [1, 3, 4]. Nevertheless, ASD prevalence has increased consistently worldwide, largely due to broader diagnostic criteria, improved case ascertainment, greater awareness, and expanded access to services [5–7]

n Brazil, the number of children receiving an ASD diagnosis through the public health system has increased in recent years. Between 2013 and 2019, the proportion of children diagnosed before four years of age also increased [8]. This trend is consistent with findings from the 2022 national census, which provided the country’s first population-based estimate of ASD, identifying approximately 2.4 million individuals (1.2% of the population) with a diagnosis [9]. Although these findings reflect important advances in early identification and recognition of ASD, they also underscore the growing demand for specialized services. Significant challenges persist, including delayed diagnosis, regional disparities, shortages of trained professionals, and limited capacity of specialized services to meet the increasing need for assessment and long-term care [10].

Evidence-based multidisciplinary rehabilitation is a cornerstone of ASD management, improving communication, adaptive functioning, cognition, and long-term quality of life, particularly when initiated during early childhood [11, 12]. In Brazil, specialized rehabilitation is primarily delivered through the Centro[1]s Especializados em Reabilitação (CER), outpatient facilities that provide diagnostic assessment, rehabilitation, multidisciplinary treatment, and assistive technology services [13]. Salvador, the capital of the state of Bahia, has a population of approximately 2.4 million inhabitants. During the study period, the city had three Ministry of Health-accredited CER II centers providing multidisciplinary rehabilitation for individuals with physical and intellectual disabilities [14]. One of these centers is the Instituto Bahiano de Reabilitação (IBR), which was accredited as a CER II in 2017 and expanded its services to provide rehabilitation for children and adults with intellectual disabilities, including ASD, in addition to physical disabilities [15].

Despite the rapid increase in ASD diagnoses and the expansion of specialized rehabilitation services, little is known about how the growing demand for ASD care has affected referral patterns, long-term follow-up, and rehabilitation service utilization in middle-income countries [15, 16]. To address this gap, we conducted a longitudinal analysis of all children receiving care at the IBR between 2017 and 2026 to (i) characterize temporal trends in ASD admissions and follow-up, (ii) evaluate changes in rehabilitation service utilization over time, and (iii) compare the duration of follow-up and retention in care. Understanding these trends is essential to inform rehabilitation service planning, workforce development, and resource allocation for the growing population of individuals with ASD [7].

## Material and Methods

### Study design and setting

We conducted a retrospective longitudinal study using routinely collected electronic health records from the IBR, a specialized multidisciplinary rehabilitation center of the *Fundação José Silveira* (FJS) in Salvador, Brazil. The IBR is an accredited Specialized Rehabilitation Center (CER II) by the Brazilian Ministry of Health and a referral center for the rehabilitation of children and adults with physical and intellectual disabilities, including autism spectrum disorder, providing comprehensive interdisciplinary care through the Brazilian Unified Health System (SUS). Located in Salvador with 2.4 million inhabitants [17], the IBR provides on the order of 90,000 appointments per year and is one of the region’s principal referral centers for developmental care in children and adolescents [18].

### Participants and eligible criteria

Our study included children and adolescents (<18 years) receiving developmental care, including services provided by the following specialties: neurology, psychiatry, physical medicine and rehabilitation (physiatry), pediatrics, psychology, occupational therapy, speech-language therapy, pedagogy, physiotherapy, physical education and nutrition. We collected information from January 2017 to July 2026. Longitudinal visit records were linked at the patient level to construct a patient-based cohort and evaluate temporal trends in new patient admissions according to study group. Children and adolescents were eligible if they attended the IBR, completed the initial intake assessment performed and attended at least two developmental care visits in a period of 3 months. Requiring a minimum of two visits ensured that participants had established clinical follow-up at the center, thereby excluding individuals who attended only for an initial screening, administrative evaluation, or referral without subsequent developmental assessment.

### Autism classification and follow-up

Three databases routinely collected at IBR were used in this study: (a) the Patient Registration Database, containing information on all outpatient visits; (b) the Pediatric Neurology Triage Database, containing data from the initial clinical assessment and diagnostic classification of patients; and (c) the Monthly Follow-up Database, comprising patients with a confirmed diagnosis of autism spectrum disorder (ASD) who were receiving multidisciplinary follow-up care.

Patients were classified into three diagnostic groups based on ICD-10 codes: (a) Childhood Autism (F84.0); (b) Other ASD, including F84.3 (other childhood disintegrative disorder), F84.5 (Asperger syndrome), F84.8 (other pervasive developmental disorders), and F84.9 (pervasive developmental disorder, unspecified); and (c) Non-ASD, comprising all remaining ICD-10 diagnoses. Patients with ICD-10 codes F84.2 (Rett syndrome) and F84.4 (overactive disorder associated with intellectual disability and stereotyped movements) were excluded from the analysis.

Follow-up indicators were derived from longitudinal visit records. Patients were classified as being under active follow-up if they had attended at least two visits during the study period and had a recorded visit within the final three months (April–June 2026). Requiring a minimum of two visits ensured that participants had established clinical follow-up at the center rather than attending a single evaluation. The last visit was defined as the most recent encounter recorded at the IBR and did not necessarily represent discharge from care. The follow-up period for each patient was calculated from the date of admission to the date of their last recorded visit at the IBR.

### Statistical analysis

Participant characteristics were summarized using descriptive statistics. Continuous variables are presented as medians and interquartile ranges (IQRs), whereas categorical variables are presented as frequencies and percentages. Comparisons across study groups (Childhood Autism, Other ASD, and Non-ASD) were performed using the Kruskal–Wallis test for continuous variables and Pearson’s χ² test for categorical variables. A subset of participants who were actively receiving care at the end of the study period were analyzed separately to characterize the population under current follow-up. The distribution of diagnostic groups among patients under active follow-up was calculated to estimate the current diagnostic composition of the IBR Developmental Care Program.

Temporal trends in both new registrations and the population under active follow-up were evaluated quarterly. For each calendar quarter, the proportion of participants in each diagnostic group was calculated and displayed using stacked bar charts to illustrate changes in the diagnostic composition over time. Absolute numbers of new registrations and patients under follow-up were also summarized by quarter. To characterize temporal trends in the annual number of new registrations and patients under follow-up, generalized additive models (GAMs) with a Poisson distribution and log link were fitted separately for each diagnostic group. Calendar year was modeled using penalized regression splines to allow for non-linear trends, and model-based predicted annual counts with 95% confidence intervals were generated for graphical visualization. The estimated smooth functions and model-predicted values were used to identify the timing of the major change in temporal trend (inflection point). A segmented Poisson regression model with a log link was subsequently fitted using the identified inflection point as the breakpoint, allowing separate annual slopes to be estimated before and after the change in trend. Annual percent changes (APC) and corresponding 95% confidence intervals (95%CI) were calculated by exponentiating the estimated regression coefficients. Because in 2026 only first-quarter data were available for the final calendar year, this year was excluded from the segmented regression analyses to avoid bias in the estimation of annual growth rates.

Follow-up duration was analyzed using Cox proportional hazards regression. Time-to-event was defined as the interval between admission and the last recorded visit (months). Models included study groups (Childhood Autism, Other ASD, and Non-ASD) and were adjusted for age at first visit, sex, and calendar year of admission. These covariates were included a priori to account for the higher proportion of males among children with ASD, differences in follow-up according to age at admission, and the shorter observation time available for more recently admitted patients. Adjusted hazard ratios (aHRs) with 95% confidence intervals (95% CIs) were estimated. Covariate-adjusted survival curves were generated from the fitted Cox model, holding continuous covariates at their median values and sex at the reference category.

Patients with unknown diagnostic classification were excluded from comparative analyses but were retained in the participant flow diagram and descriptive figures illustrating cohort derivation (Fig 1). Statistical significance was defined as a two-sided P < 0.05. All analyses were performed using R version 4.5.1.

**Fig 1.**
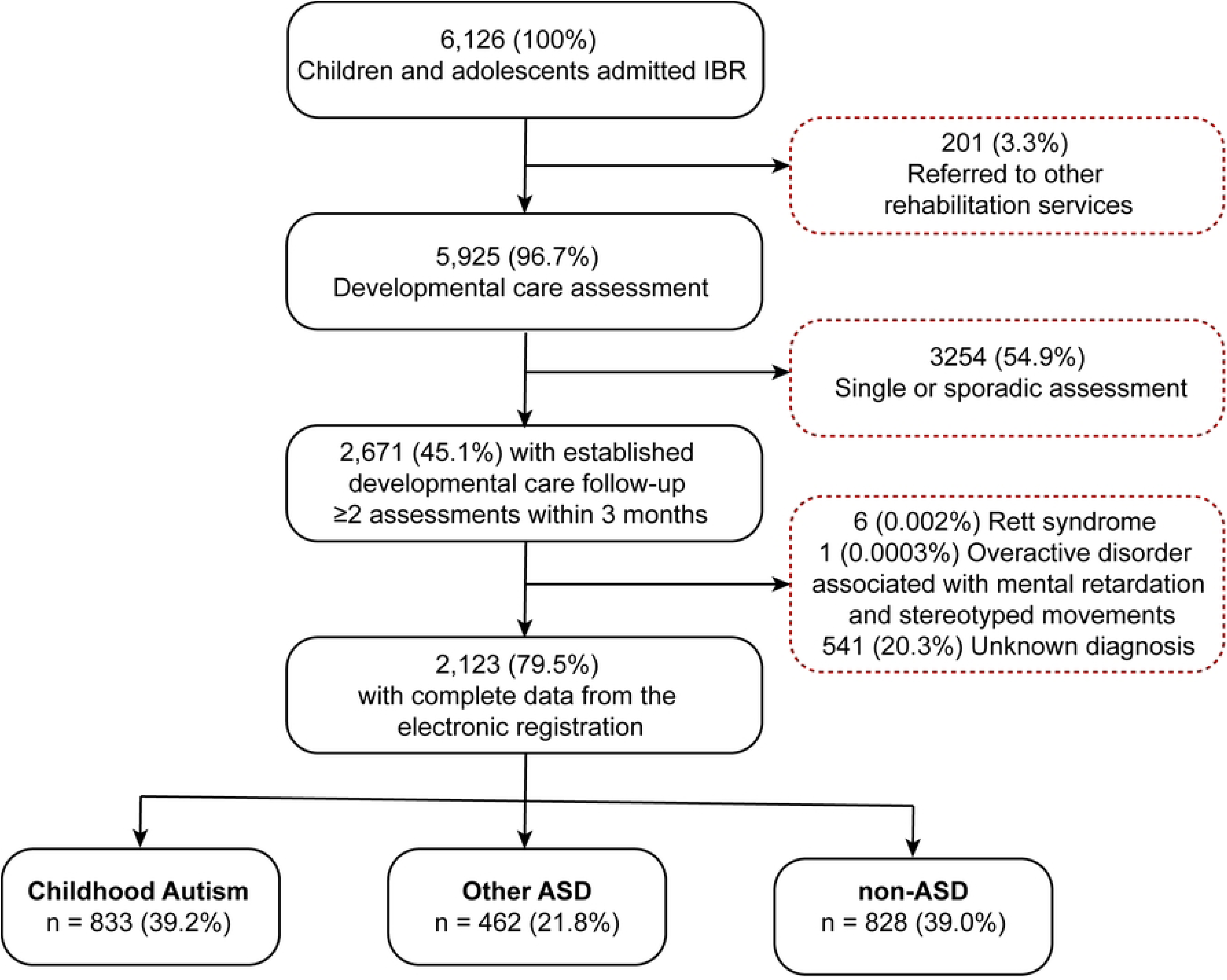
Flowchart.

### Ethic statement

This study was approved by the Research Ethics Committee of the Maternidade Climério de Oliveira, Federal University of Bahia (UFBA) under the project *“Clinical and epidemiological profile of patients attending a Specialized Rehabilitation Center II (CER II) in Salvador, Bahia”* (CAAE: 90223225.7.0000.5543; approval no. 7.719.876). As this was a retrospective study based on routinely collected clinical data, the requirement for informed consent was waived by the Ethics Committee.

## Results

Of the 6,126 children and adolescents admitted to IBR during the study period, 2,123 met the eligibility criteria and were included in the analysis. Participants were classified into three study groups: Childhood Autism (n = 833, 39.2%), Other ASD (n = 462, 21.8%), and non-ASD diagnoses (n = 828, 39.0%). The main characteristics of the participants are summarized in Table 1. Briefly, children with Childhood Autism were admitted at a younger age than those with non-ASD diagnoses (median age at first visit: 4 vs. 5 years), whereas children with Other ASD were admitted at the youngest age (median: 2 years). Childhood Autism was more common among males (78%). Patients with Childhood Autism and Other ASD had a higher number of visits and longer follow-up than those with non-ASD diagnoses. At the end of the study period, 608 (28.5%) patients were actively receiving follow-up care, including 54% of those with Childhood Autism, 18% of those with Other ASD, and 9.7% of those with non-ASD diagnoses. Among patients who remained under active follow-up, current age did not differ significantly between groups. However, patients with Childhood Autism, and particularly those with Other ASD, had accumulated more visits and longer follow-up than patients with non-ASD diagnoses (Table 1). The non-ASD diagnoses group included patients with a broad spectrum of referral diagnoses (Table S1).

**Table 1.**
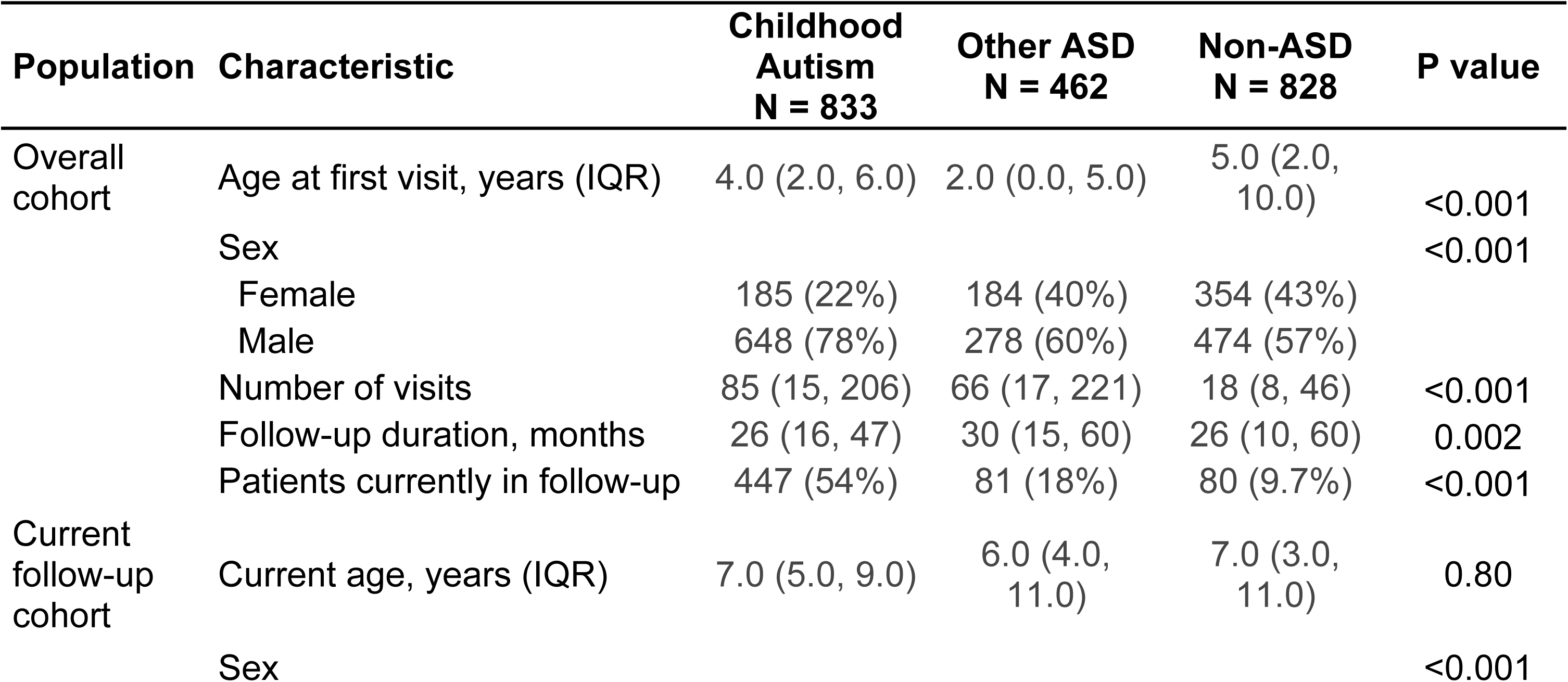
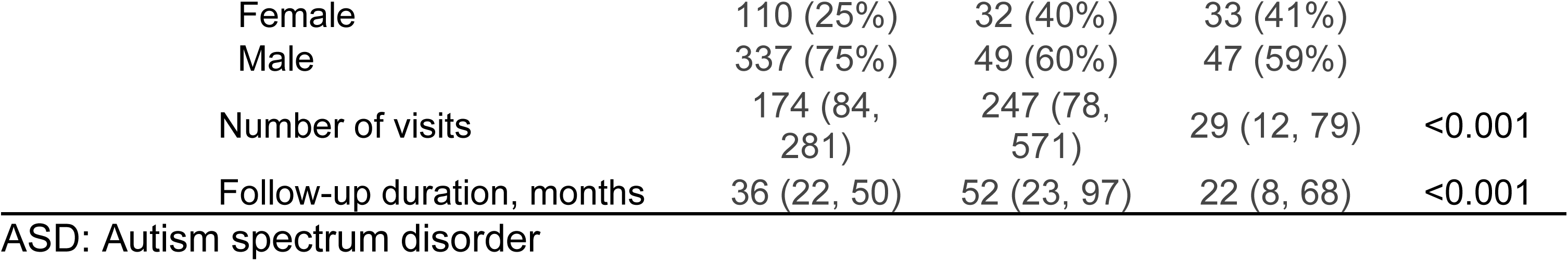
Participant characteristics of the overall cohort and the current follow-up cohort at IBR.

| Population | Characteristic | Childhood Autism<br>N = 833 | Other ASD<br>N = 462 | Non-ASD<br>N = 828 | P value |
| --- | --- | --- | --- | --- | --- |
| Overall cohort | Age at first visit, years (IQR) | 4.0 (2.0, 6.0) | 2.0 (0.0, 5.0) | 5.0 (2.0, 10.0) | <0.001 |
|  | Sex |  |  |  | <0.001 |
|  | Female | 185 (22%) | 184 (40%) | 354 (43%) |  |
|  | Male | 648 (78%) | 278 (60%) | 474 (57%) |  |
|  | Number of visits | 85 (15, 206) | 66 (17, 221) | 18 (8, 46) | <0.001 |
|  | Follow-up duration, months | 26 (16, 47) | 30 (15, 60) | 26 (10, 60) | 0.002 |
|  | Patients currently in follow-up | 447 (54%) | 81 (18%) | 80 (9.7%) | <0.001 |
| Current follow-up cohort | Current age, years (IQR) | 7.0 (5.0, 9.0) | 6.0 (4.0, 11.0) | 7.0 (3.0, 11.0) | 0.80 |
|  | Sex |  |  |  | <0.001 |

|  |  |  |  |  |
| --- | --- | --- | --- | --- |
| Female | 110 (25%) | 32 (40%) | 33 (41%) |  |
| Male | 337 (75%) | 49 (60%) | 47 (59%) |  |
| Number of visits | 174 (84, 281) | 247 (78, 571) | 29 (12, 79) | <0.001 |
| Follow-up duration, months | 36 (22, 50) | 52 (23, 97) | 22 (8, 68) | <0.001 |

ASD: Autism spectrum disorder

The age distribution at admission differed across diagnostic groups. Children with Childhood Autism were most commonly admitted between 3 and 5 years of age and exhibited a marked male predominance across all age groups. Children with Other ASD were admitted at younger ages than those with non-ASD diagnoses and showed a more balanced sex distribution. In contrast, children with non-ASD diagnoses were admitted across a broader age range, extending into later childhood and adolescence (Fig. 2A). By the last visit, the age distribution had shifted toward older age groups, particularly among children with Childhood Autism, reflecting continued follow-up after admission (Fig. 2B). Consistent with this pattern, children with Childhood Autism had the longest follow-up duration, whereas those with non-ASD diagnoses were predominantly followed for shorter periods, although a subset remained under long-term care. (Fig. 2C).

**Fig 2.**
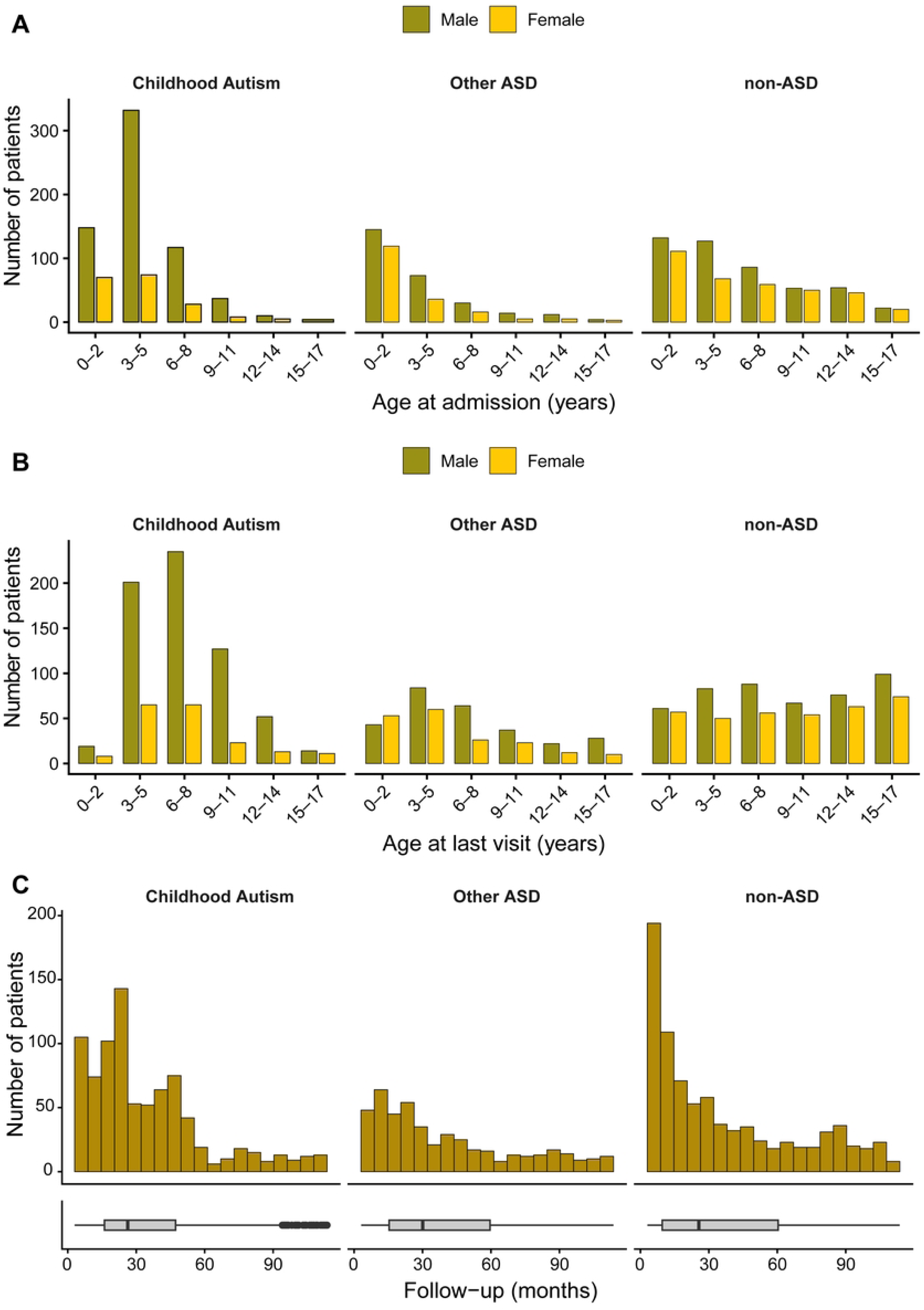
Demographic characteristics and follow-up duration of patients. (A) Distribution of patients by age at admission (3-year age groups) and sex for patients with Childhood Autism, Other ASD, and non-ASD diagnoses. Bars represent the number of patients within each age category, stratified by sex. (B) Distribution of patients by age at the last follow-up visit (3-year age groups) and sex for the same study groups. Bars represent the number of patients within each age category, stratified by sex. (C) Distribution of follow-up duration after admission. Histograms show the frequency distribution of follow-up time (months), and boxplots summarize the median, interquartile range (IQR), and range of follow-up duration.

During the study period, the profile of new patient admissions at IBR changed substantially (Fig. 3A and Table S2). At the beginning of the study, children with Childhood Autism accounted for only 7.5% (29/389) of all new admissions, whereas children with non-ASD diagnoses represented 58.6% (228/389). Over time, the proportion of new admissions for Childhood Autism increased steadily, exceeding 50% from 2022 onward and reaching 88.2% (127/144) in the third quarter of 2024. In contrast, the proportion of new admissions for children with non-ASD diagnoses declined sharply, from 58.6% to 2.1%, while admissions for Other ASD remained relatively stable throughout the study period (Fig. 3A and Table S2). The marked reduction in admissions observed in 2020 (Fig. 3A) coincided with the COVID-19 pandemic, reflecting the widespread reorganization of healthcare services and clinical protocols in response to the public health emergency [19]. Consistent with these quarterly trends, generalized additive models (GAMs) showed a marked increase in annual admissions for Childhood Autism through 2023, a progressive decline in admissions for non-ASD diagnoses throughout the study period, and relatively modest variation in admissions for Other ASD (Fig. 3C and Table S3).

**Fig 3.**
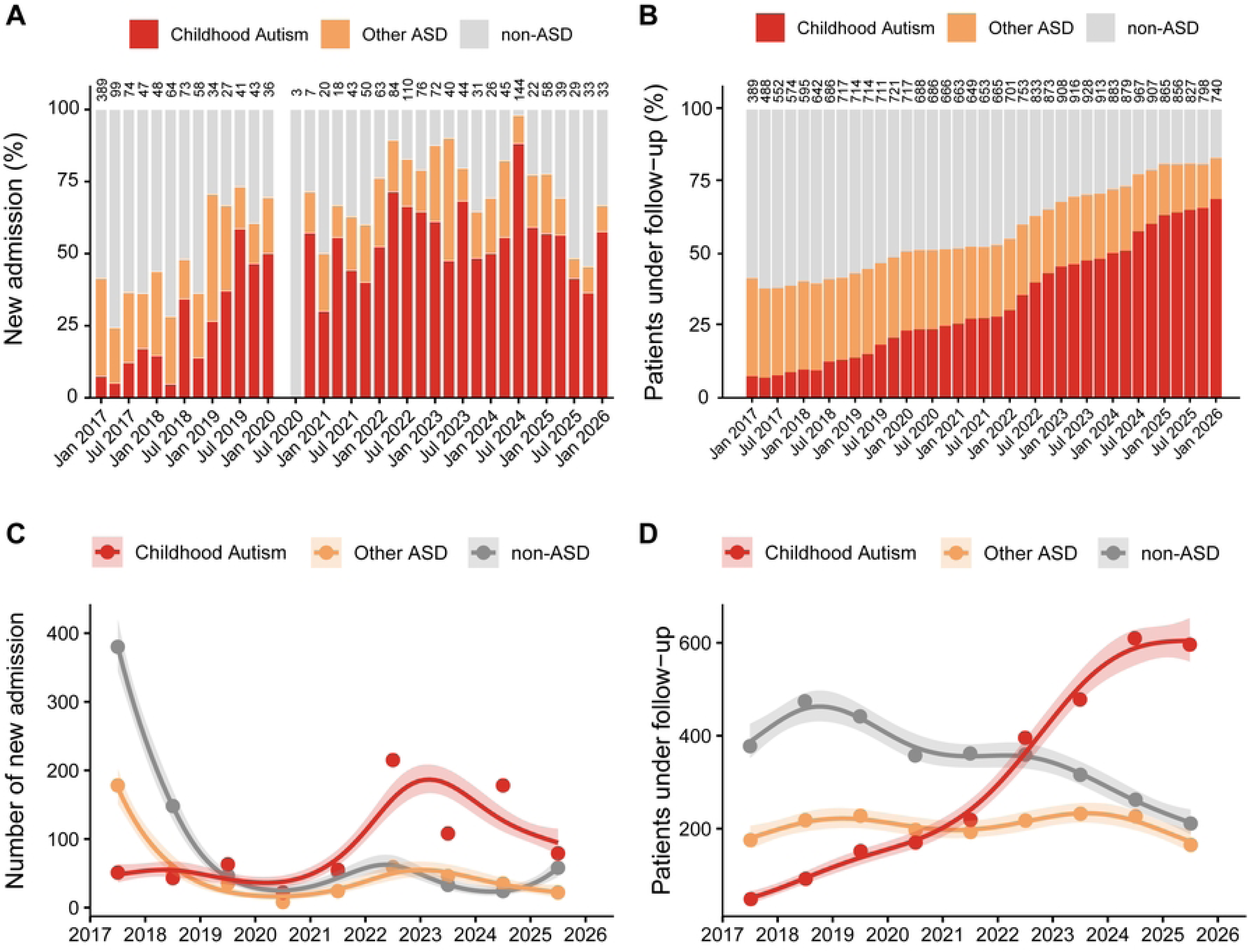
Temporal changes in patient admissions and follow-up at IBR, 2017–2026. (A) Quarterly distribution of patient admissions according to diagnostic group (Childhood Autism, Other ASD, and non-ASD diagnoses). Numbers above the bars indicate the total number of admissions per quarter. (B) Quarterly distribution of patients under follow-up according to diagnostic group. (C) Annual observed numbers of patient admissions (points) and generalized additive model (GAM)-estimated temporal trends (solid lines) with 95% confidence intervals for each diagnostic group. (D) Annual observed numbers of patients under follow-up (points) and GAM-estimated temporal trends (solid lines) with 95% confidence intervals for each diagnostic group.

The increase in Childhood Autism admissions was accompanied by a substantial expansion in the number of children receiving rehabilitation services at IBR. The number of children with Childhood Autism under follow-up increased from 29 in the first quarter of 2017 to a peak of 558 in the third quarter of 2024, remaining above 500 through the end of the study period (Fig. 3B,D and Table S3). In contrast, the number of children with non-ASD diagnoses under follow-up declined steadily from 228 to 125, while the number of children with Other ASD remained relatively stable before gradually decreasing after 2024. Consequently, the clinical workload at IBR shifted progressively from being dominated by children with non-ASD diagnoses to predominantly serving children with Childhood Autism (Fig. 3B,D).

Segmented Poisson regression identified significant changes in temporal trends after 2023 (Table 2). Between 2017 and 2022, admissions of children with Childhood Autism increased by 30.2% annually (95% CI, 24.7–35.9; *P*<0.001). After 2023, this trend reversed, with admissions declining by 19.6% annually (95% CI, −27.9 to −10.2; *P*<0.001). Admissions of children with Other ASD declined before 2023 (APC, −21.7%; 95% CI, −25.6 to −17.6; *P*<0.001) but showed no significant change thereafter (APC, 3.6%; 95% CI, −16.5 to 28.4; *P*=0.262). In contrast, admissions of children with non-ASD diagnoses decreased sharply before 2023 (APC, −39.1%; 95% CI, −41.8 to −36.3; *P*<0.001), followed by a marked increase after 2023 (APC, 92.0%; 95% CI, 61.2–129.0; *P*<0.001). Despite this relative increase, the absolute number of admissions for children with non-ASD diagnoses remained substantially lower than at the beginning of the study period (Fig. 3A and Table S2). Trends among patients under follow-up differed from those observed for admissions. Although admissions of children with Childhood Autism declined after 2023, the number of children receiving ongoing rehabilitation continued to increase, albeit at a slower rate, from an annual increase of 41.8% (95% CI, 38.0–45.8; *P*<0.001) before 2023 to 9.9% (95% CI, 4.3–15.8; *P*<0.001) thereafter. In contrast, the number of children with Other ASD under follow-up increased slightly before 2023 (APC, 2.7%; 95% CI, 0.1–5.2; *P*=0.039) but declined after 2023 (APC, −12.3%; 95% CI, −19.4 to −4.6; *P*=0.002). Meanwhile, the number of children with non-ASD diagnoses under follow-up declined throughout the study period, with the rate of decline accelerating after 2023 (APC, −4.7% before 2023 vs. −20.8% after 2023). Consequently, children with Childhood Autism represented an increasingly large proportion of the active rehabilitation caseload despite the decline in new admissions after 2023.

**Table 2.**
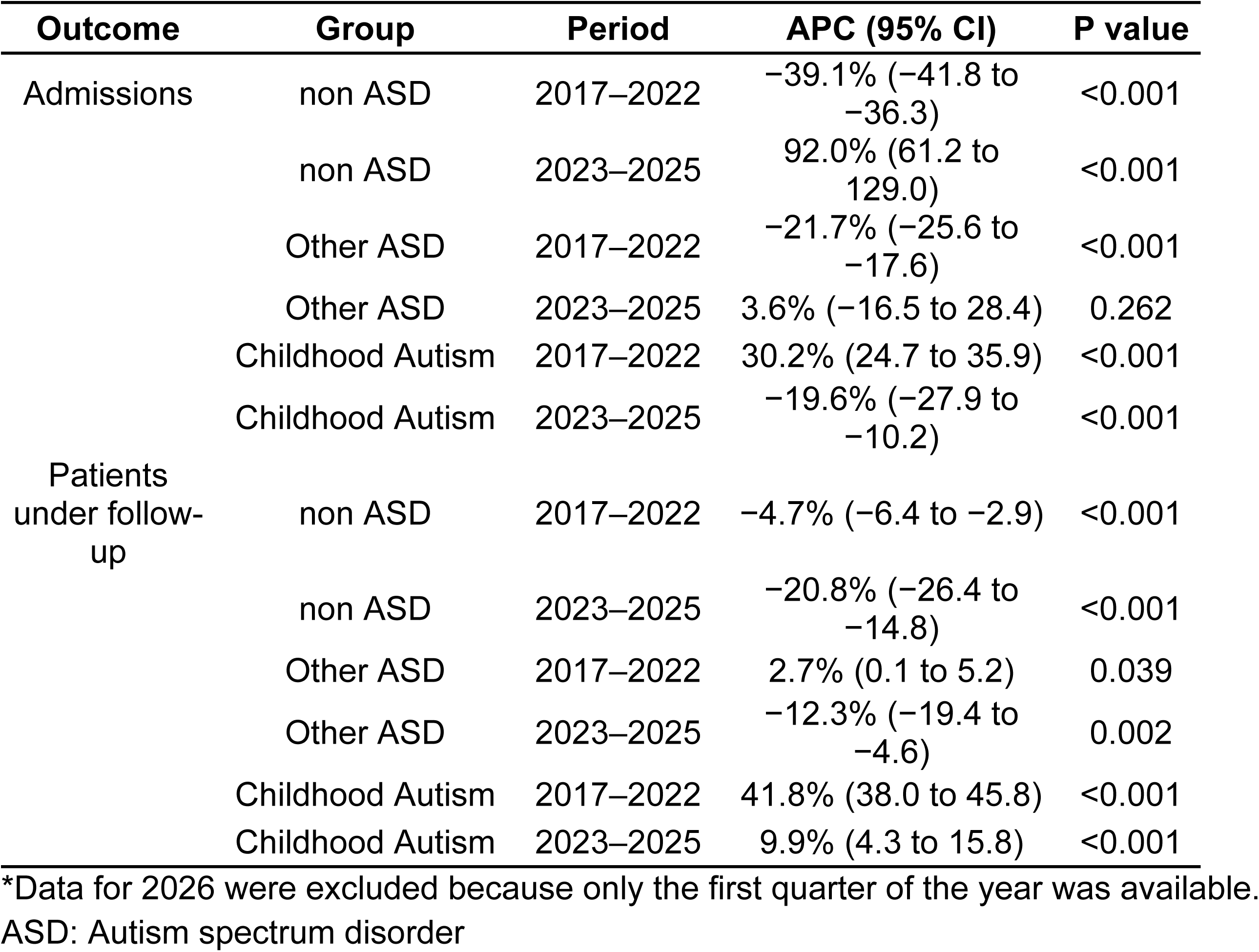
Annual percent change (APC) in patient admissions and patients under follow-up before and after 2023, estimated using segmented Poisson regression.

After adjustment for age at first visit, sex, and calendar year of admission, children with Childhood Autism remained under follow-up at IBR substantially longer than those with non-ASD diagnoses (Fig. 4 and Table 3). Compared with children with non-ASD diagnoses, those with Childhood Autism had a 65% lower hazard of reaching their last recorded follow-up visit (adjusted hazard ratio [aHR], 0.35; 95% CI, 0.30–0.40; P<0.001), whereas children with Other ASD had a 21% lower hazard (aHR, 0.79; 95% CI, 0.69–0.89; P<0.001). Older age at first visit was associated with a modestly higher hazard of reaching the last recorded follow-up visit (aHR per year, 1.03; 95% CI, 1.02–1.05; P<0.001), whereas sex was not associated with follow-up duration (aHR for males, 0.97; 95% CI, 0.87–1.08; P=0.549). More recent calendar year of admission was associated with a higher hazard of reaching the last recorded follow-up visit (aHR per calendar year, 1.22; 95% CI, 1.19–1.25; P<0.001).

**Fig 4.**
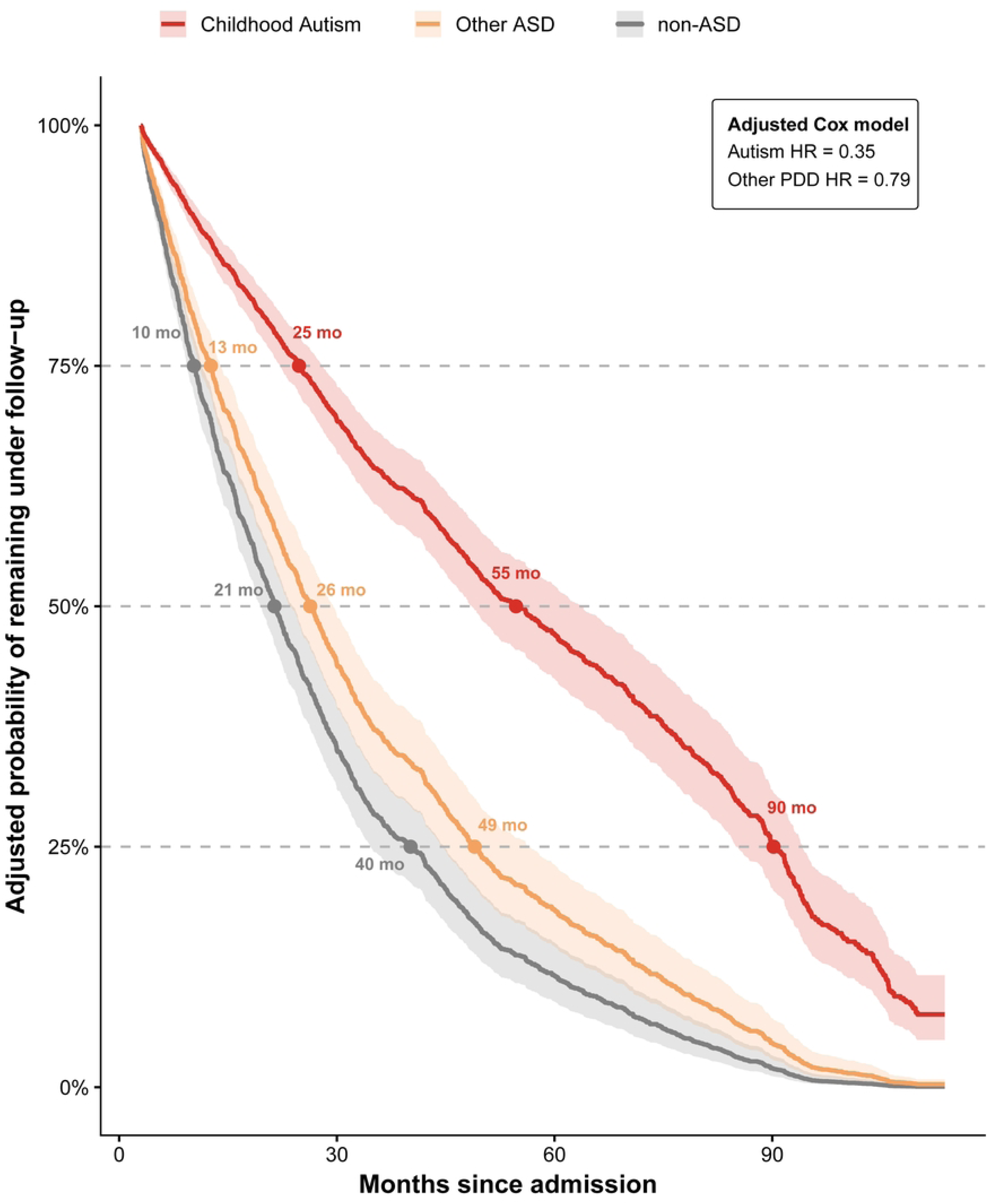
Adjusted probability of remaining under follow-up according to study groups. Covariate-adjusted survival curves derived from a Cox proportional hazards model comparing children with Childhood Autism, Other ASD, and non-ASD diagnoses. The model was adjusted for age at first visit, sex, and calendar year of admission. Shaded areas represent 95% confidence intervals.

**Table 3.** Multivariable Cox proportional hazards model of time to last recorded follow-up visit.

| Predictor | Adjusted HR (95% CI) | P value |
| --- | --- | --- |
| Study group |  |  |
| Non-ASD | Reference | — |
| Other ASD | 0.79 (0.69–0.89) | $<0.001$ |
| Childhood Autism | 0.35 (0.30–0.40) | $<0.001$ |
| Age at first visit (per year) | 1.03 (1.02–1.05) | $<0.001$ |
| Male sex | 0.97 (0.87–1.08) | 0.549 |
| Calendar year of admission | 1.22 (1.19–1.25) | $<0.001$ |

HR, hazard ratio; CI, confidence interval; ASD, autism spectrum disorder. Hazard ratios (HRs) and 95% confidence intervals (CIs). The event was the last recorded follow-up visit, with lower HRs indicating longer retention under follow-up.

## Discussion

In this longitudinal study of a large rehabilitation center for intellectual and physical disabilities in Salvador, Brazil, we observed a profound shift in the profile of children receiving developmental care over the last decade. Childhood Autism evolved from representing a small minority of patients in 2017 to becoming the predominant condition managed by the service. This transition was driven by a sustained increase in admissions through 2023, together with the substantially longer duration of follow-up among children with Childhood Autism compared with those with non-ASD diagnoses. As a result, the number of children receiving ongoing rehabilitation for Childhood Autism continued to increase despite the subsequent decline in new admissions, reflecting the cumulative demand generated by prolonged rehabilitation needs. Consequently, the growing occupancy of rehabilitation services by children with Childhood Autism was accompanied by a progressive reduction in the proportion of patients with non-ASD diagnoses receiving care. Together, these findings suggest that the increasing demand for long-term autism rehabilitation has fundamentally reshaped the organization of rehabilitation services and may require structural adaptations in service capacity and resource allocation to ensure equitable access for children with a broad range of developmental conditions.

Previous studies have shown that the increasing prevalence of autism has placed growing pressure on healthcare systems, with demand for diagnostic and treatment services outpacing the capacity of many specialized centers. Autism affects approximately 1% of the global population and has become an increasingly important public health challenge because of the marked rise in reported diagnoses over the past two decades [20, 21]. Although part of this increase is likely attributable to greater awareness, broader diagnostic criteria, improved case ascertainment, and a decline in the use of alternative developmental diagnoses, several studies suggest that these factors do not fully explain the observed trends [22, 23]. Consequently, the demand for autism-related health services has increased worldwide, placing growing pressure on healthcare systems [7]. Evidence from low- and middle-income countries remains limited, however, because of the scarcity of population-based surveillance and health service studies. In Brazil, the 2022 national census estimated that approximately 1.2% of the population (2.4 million individuals) had received an autism diagnosis, underscoring autism as an emerging public health priority. Recent studies have also highlighted substantial unmet healthcare needs and persistent barriers to accessing services for Brazilian children with autism [24]. Our findings complement this evidence by identifying a potential mechanism underlying these challenges in a middle-income country. Although admissions of children with Childhood Autism increased during the study period, these children entered rehabilitation at younger ages and remained under follow-up substantially longer than those with non-ASD diagnoses, resulting in the progressive accumulation of patients requiring ongoing rehabilitation. This pattern is consistent with recent frameworks describing autism as a lifelong condition with evolving support needs across the life course [25]. Therefore, individuals with autism use healthcare services more intensively than the general population, highlighting the importance of considering long-term service demand when planning healthcare resources [26]. Consequently, autism became the predominant condition in the rehabilitation caseload despite a subsequent decline in new registrations, suggesting that increasing long-term service occupancy may progressively reduce capacity for new referrals and other developmental conditions. These findings highlight the need to adapt rehabilitation services, workforce capacity, and resource allocation to meet the growing demand for autism care while maintaining equitable access for all children with developmental disabilities.

In our study, males predominated among patients with autism spectrum disorder (ASD), in line with the established literature [27]. However, this sex imbalance should be interpreted cautiously, because the observed male excess may not fully represent true etiologic prevalence. Evidence suggests that girls and women with ASD are more likely to be under-recognized, diagnosed later, or missed altogether, partly because of sex-related differences in clinical presentation and limitations of diagnostic tools developed largely from male samples [27, 28]. A systematic review and meta-analysis reported that the commonly cited 4:1 male-to-female ratio is likely an overestimate, with a pooled ratio closer to 3:1 in higher-quality studies and in population-screened samples, supporting the presence of diagnostic gender bias [27]. Consistently, registry-based data show that the male predominance is smaller in adults than in children, suggesting that delayed identification in females may contribute to the apparent sex difference observed in clinical settings [28]. Therefore, our findings reinforce the need for sex-sensitive diagnostic approaches to improve recognition of ASD in females and to better estimate the true sex distribution of the disorder.

## Limitation

This study has some limitations. First, diagnostic classification was based on routinely collected electronic health records and ICD-10 codes assigned during routine clinical care rather than standardized research assessments. As a tertiary rehabilitation center, IBR receives patients with an established referral diagnosis and does not routinely reassess the diagnosis for research purposes. Diagnostic reclassification was uncommon and, when performed, was generally related to refining the autism subtype or severity rather than changing the diagnosis itself. Nevertheless, follow-up was established by multidisciplinary clinical teams and integrated across multiple institutional databases, reflecting real-world clinical practice. Second, participants with undefined diagnostic information were excluded from the comparative analyses, which may have introduced selection bias. However, sensitivity analyses indicated that most excluded patients had Non-ASD conditions and only sporadic contact with the rehabilitation service, suggesting that their exclusion is unlikely to have materially affected the main findings (Table S4). Finally, the analysis was restricted to routinely collected data available from 2017 onward and did not include information on waiting times, referral pathways, treatment intensity, or clinical outcomes. Nevertheless, the long observation period and comprehensive longitudinal follow-up provided a unique opportunity to characterize how increasing demand for autism care has reshaped rehabilitation service utilization in a middle-income country.

## Conclusion

The rapid increase in autism diagnoses has fundamentally reshaped rehabilitation service utilization at a large public referral center in Brazil. Because children with autism entered care earlier and remained under follow-up longer, they progressively became the predominant users of rehabilitation services. These findings suggest that referral trends alone do not adequately capture future demand for rehabilitation services. Planning for autism care should therefore account for long-term service utilization and patient retention, alongside changes in disease prevalence, to ensure sufficient capacity and equitable access for children with autism and other developmental disabilities.

## Data Availability

Data Availability: The R script used for all analyses is publicly available on GitHub at https://github.com/pkjpablo/Autismo. The anonymized dataset supporting the findings of this study is available in the same repository.

## List of supporting information captions

Table S1. Referral diagnoses recorded among patients in the Non-ASD group

Table S2. Quarterly numbers of new registrations and patients under follow-up, and the proportion of patients in each patient group, 2017–2026.

Table S3. Annual observed counts and GAM-estimated counts (95% confidence intervals) for new registrations and patients under follow-up according to diagnostic group, 2017–2026.

Table S4. Comparison of participants included in the analysis and those with missing data.

